# COVID 19, Consumption and Inequality: A Systematic Analysis of Rural Population of India

**DOI:** 10.1101/2021.06.08.21258525

**Authors:** Mudit Kapoor, Shamika Ravi, A.K. Shiva Kumar

## Abstract

**Background:** COVID19 pandemic has had major impact on consumption levels and inequality within India. Government policy interventions have targeted poor households for cash and food transfers. It is important, however, to study the impact of the pandemic on consumption levels of non-poor in India, and in particular the middle class. In this paper, we aim to quantify the changes in consumption levels and inequality over time, across all groups of rural households in India.

**Methods:** We analyze three rounds of COVID 19-related shock surveys between May and September 2020. These surveys cover rural households of six large states in India and are representative of more than 442 million (52% of India’s rural population).

**Findings:** In the early phase of the pandemic, it was the bottom 40% of households that experienced the most severe decline in consumption. But as the pandemic deepened, consumption declined across all classes of households. Besides the poorest, it was particularly severe for the middle class (defined as 40%-80%). We also measure consumption inequality over time and find that the Gini coefficient of consumption distribution increased significantly.

**Interpretation:** In addition to focusing on poor households, policy responses to alleviate people’s sufferings would have to consider a more comprehensive boost to consumption and compensate for the reduced consumption among middle class families as well.

**Funding:** None.

## 1. Introduction

This paper analyzes the impact of the COVID 19 pandemic on consumption and inequality across different classes of people - the rich, middle class, and the poor - in rural India. The impact of the lockdowns during the pandemic (ever since it was first imposed on March 25, 2020) has been extremely severe on the overall economy and particularly on consumption. For example, the Index of Industrial Production (IIP) for consumer durables fell drastically from 117 in February 2020 to 6 in April 2020 when India was in the midst of a national lockdown. The IIP for non-durable goods fell from 153 to 73 during the same period.^1^ Even though this is informative in terms of overall contraction in the economy, it provides little guidance in framing targeted policies towards different classes of people affected by the pandemic.

As of 17 May 2021, India has had more than 25 million COVID 19 cases, the second-highest in the world, and is in the middle of a massive second wave of infections, with more than three times the number of daily cases as compared to the first wave that started in March 2020 and peaked in September 2020.^2^ This has compelled several state governments to impose micro lockdowns to contain the spread of the virus. The magnitude and the suddenness of the second wave has created an atmosphere of uncertainty, fear, and panic with serious negative implications for economic recovery.^3,4^ To alleviate the economic fallout of the COVID 19 pandemic, the central and state governments have undertaken unprecedented fiscal expansion, such as cash and kind transfers targeted at the poor, liquidity support to especially to small and medium enterprises to prevent layoffs and defaults, and increased infrastructure spending to deal with the first wave of the pandemic.^5^ The enormous magnitude of the second wave has once again prompted discussions of another round of fiscal expansion.

This paper examines the adequacy of the conventional focus of alleviating the suffering of the poor during such a humanitarian crisis by examining the impact of the COVID-19 on the non-poor as well. We do this by quantifying the decline in consumption across different classes and identifying the extent of losses suffered by non-poor income groups as well. Such an analysis is necessary to guide policymakers to frame policies in accordance with the needs of different classes of people. Our paper complements earlier research on the socio-economic impact of COVID 19 that has focused on income losses across different occupational groups, cultivator, business, salaried, and casual workers, in urban areas.^6^ Instead of income, our focus is on consumption and consumption inequality across varying classes, namely the rich, middle class, and the poor in rural areas.

## 2. Data

We use the publicly available World Bank’s COVID 19 related shocks survey conducted between May and September 2020 in three rounds from rural areas of six large states in India: Andhra Pradesh, Bihar, Jharkhand, Madhya Pradesh, Rajasthan, and Uttar Pradesh.^7^ The sample is representative of more than 442 million (52% of India’s rural population) rural inhabitants in these six states.^7^ Round 1 was conducted between May 5-10, 2020; Round 2 was conducted between July 19-23 2020; and Round 3 was conducted between September 20-24, 2020. The survey covered the following main issues: (a) Agriculture, such as price realizations, acreage decision, access to inputs, and credits, etc. (b) Income and Consumption, for example, changes in wage rates, job duration, household consumption expenditure in the week prior to the survey. In Round 1 (conducted in May 2020), the survey also asked about the household consumption expenditure in the month of February 2020. (c) Migration, rates of in-migration, employment status, and plans of return migration. (d) Access to relief in the form cash or kind, quantities of relief, and constraints in access to relief. (f) Access to health facilities, foregone healthcare, knowledge of COVID 19 related symptoms and protective behaviors.

The survey was conducted using Computer Assisted Telephone Interview techniques, and the software used was SurveyCTO. The sample size for Round 1 was 4,550 and the response rate was close to 55%, for Round 2 the sample size was 5,005 and the response rate was approximately 46%, and for Round 3 the sample size was 5,200 and the response rate was close to 55%. However, not all households that were surveyed responded to the consumption questions in the survey: 3,789 (83%) households responded in Round 1, we have consumption response from households, 4,176 (83%) households responded in Round 2, and 4,422 (85%) of the households responded to the consumption survey in Round 3.

For this paper, we use data on consumption. In order to make the estimates comparable across the states and to create pooled estimates across the six states, weights were applied to the information provided by the sampled households. Details on the data, questionnaire, and the sampling methodology are available on the World Bank’s website.^7^

## 3. Results

The findings of the survey are summarized below:

Age of the respondents: The average age of respondent in Round 1 was 37.5 years (95% CI: 36.4 to 38.5), in Round 2 it was 41.3 years (95% CI: 40.8 to 41.9), while in Round 3 it was 40.7 years (95% CI: 40.1 to 41.4).

Household characteristics: The average household size in Rounds 1, 2, and 3 was 6.6 (95% CI: 6.3 to 6.8), 6.0 (95% CI: 5.7 to 6.3), and 5.4 (95% CI: 4.4 to 6.5), respectively. In Round 1, proportion of male headed households was 85% (95% CI: 82.0% to 87.5%), in Round 2 it was 84.1% (95% CI: 81.6% to 86.3%), while in Round 3 it was 84.6% (95% CI: 82% to 86.8%). Detailed results are available in Table 1 and Figure 1.

**Table 1:**
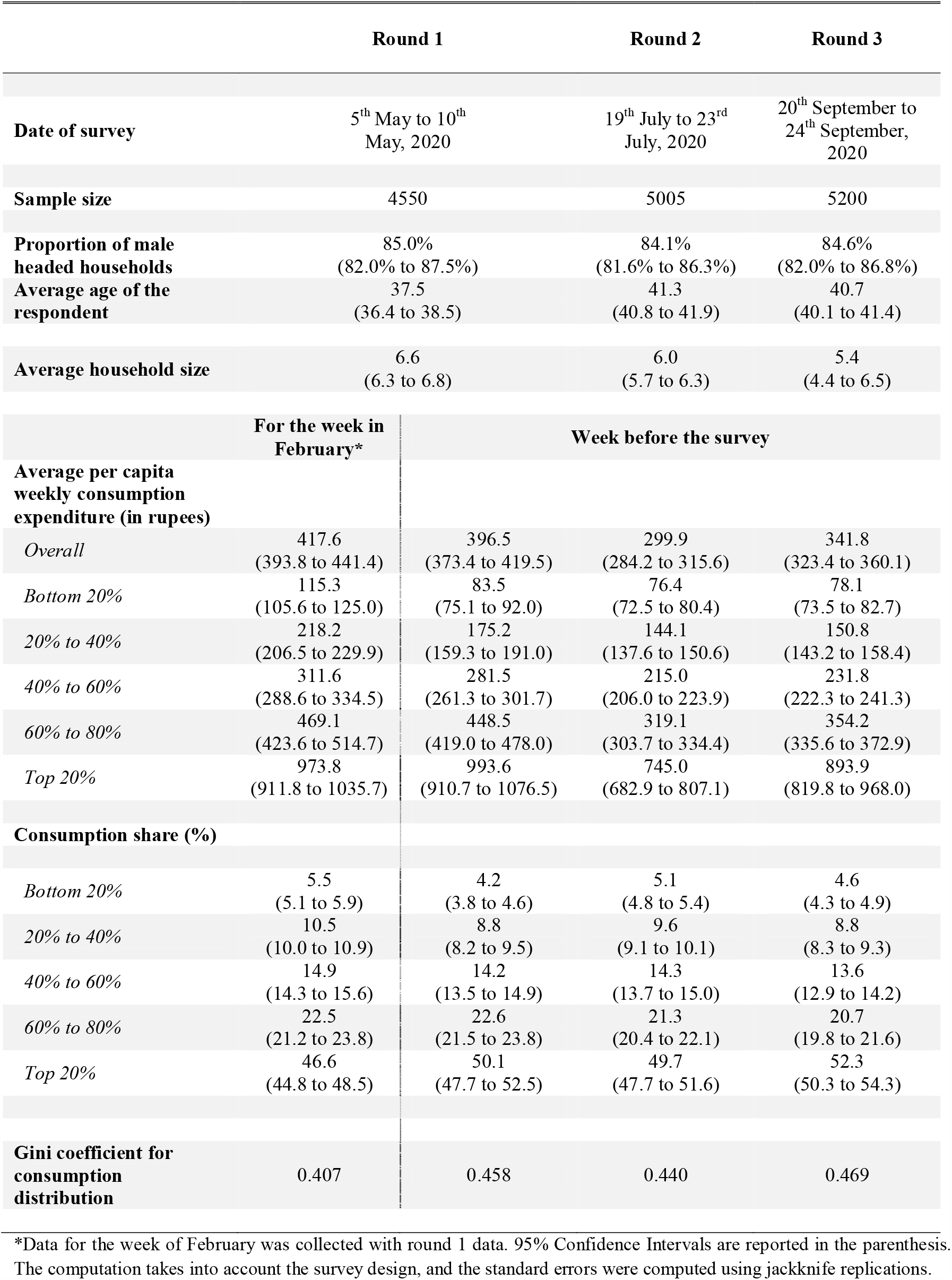
Summary table

**Figure 1:**
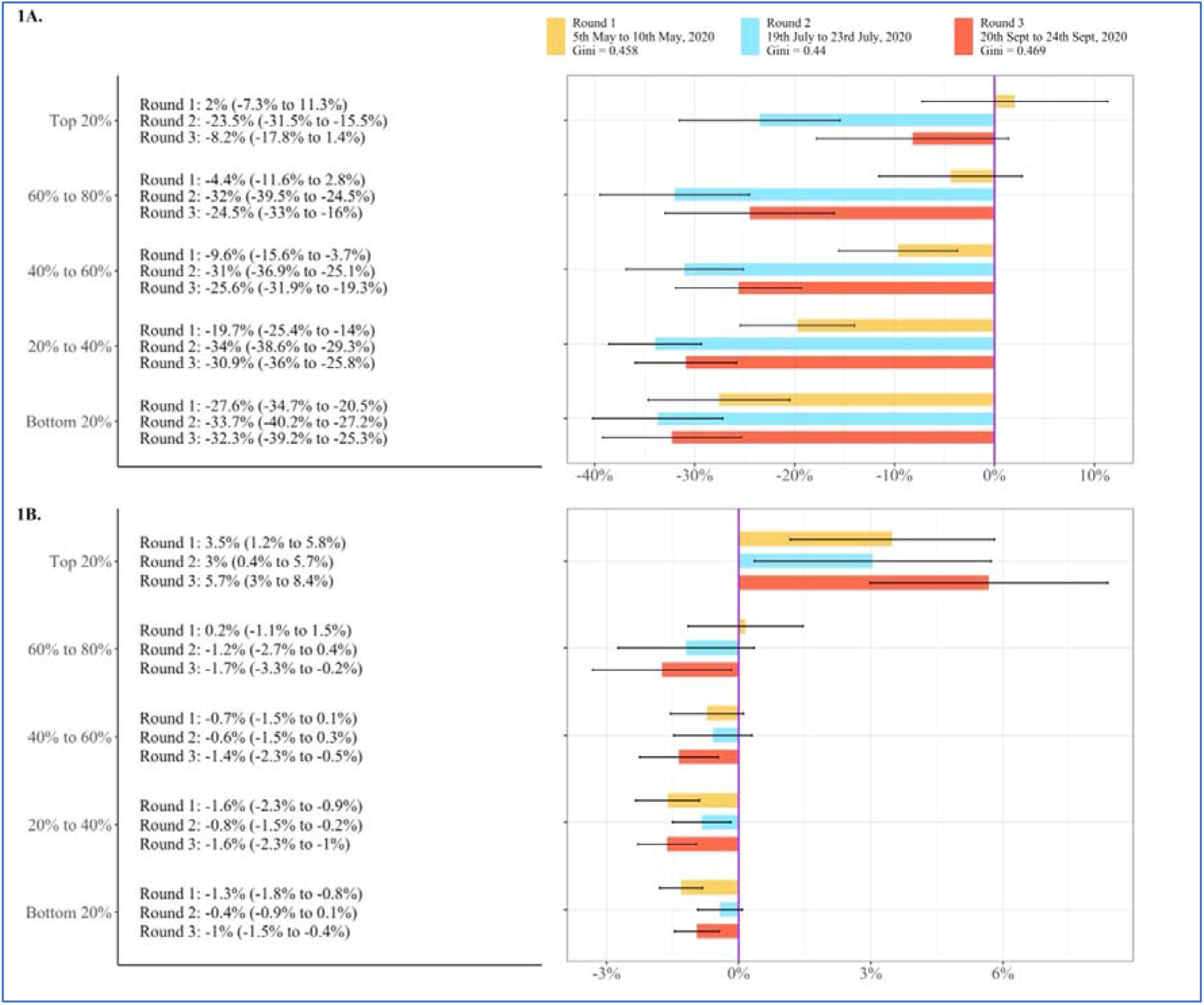
1A. Percentage change in average per capita weekly consumption expenditure for quintiles & 1B. Change in consumption share (%), as compared to week in February. 95% Confidence Intervals are reported in the parenthesis. The computation takes into account the survey design, and the standard errors were computed using jackknife replications.

Household expenditures: Overall, the weekly per capita household expenditure declined from INR 417.6 (95% CI: INR 393.8 to 441.4) in the week of February 2020 to 396.5 (95% CI: INR 373.4 to 419.5) in Round 1 (which was the week prior to May 5-10, 2020). It declined further to INR 299.9 (95% CI: INR 284.2 to 315.6) in Round 2 (which was week prior to July 19-23, 2020). The weekly per capita household expenditure recovered in Round 3 (which was week prior to September 20-24, 2020) from Round 2, to INR 341.8 (95% CI: INR 323.4 to 360.1). However, this change in weekly per capita household expenditure was not uniform across the quintile classes, and across the rounds (see table 1).

The decline in weekly per capita household expenditure in Round 1 was the highest in percentage terms for the bottom 20%, where it fell from INR 115.3 in the week of February to INR 83.5 (95% CI: INR 75.1 to 92.0) - a decline of 27.6% (95% CI: -34.7% to -20.5%). This was followed by the decline in consumption of bottom 20% to 40%, where consumption fell from INR 218.2 (95% CI: INR 206.5 to 229.9) to INR 175.2 (95% CI: INR 159.3 to 191.0) - a decline of 19.7% (95% CI: -25.4% to -14%). For the quintile class 40% to 60%, during the same period, consumption declined by 9.6% (95% CI: -15.6% to -3.7%) and fell from INR 311.6 (95% CI: INR 288.6 to 334.5) to INR 281.5 (95% CI: INR 261.3 to 301.7). However, for the 60% to 80% quintile class, during the same period, consumption declined insignificantly by 4.4% (95% CI: -11.6% to 2.8%) and fell from INR 469.1 (95% CI: INR 423.6 to 514.7) to INR 448.5 (95% CI: INR 419.0 to 478.0). In contrast, for the top 20%, consumption increased marginally though insignificantly by 2% (95% CI: -7.3% to 11.3%), from INR 973.8 (911.8 to 1,035.7) to INR 993.6 (910.7 to 1,076.5). These results suggest that in the early periods of the pandemic, and during the lockdown, consumption decline was more severe for the bottom 40% compared to the top 40% which experienced an insignificant change in consumption.

As the pandemic deepened, however, there was a severe contraction in consumption across all quintile classes. For example, when we compare Round 2 consumption (in July 2020) to the week of February 2020, we find that that consumption decline is approximately the same for the bottom 20% as compared to Round 1 - at -33.7% (95% CI: -40.2% to -27.2%). For the quintile class 20% to 40%, per capita weekly consumption expenditure declined further by 34% (95% CI: -38.6% to 29.3%) to INR 144.1 (95% CI: INR 137.6 to 150.6). Even for the top 20%, the weekly per capita expenditure declined in Round 2 as compared to the week of February, by -23.5% (95% CI: -31.5% to -15.5%) - a decrease from INR 973.8 (95% CI: INR 911.8 to 1035.7) to INR 745.0 (95% CI: INR 682.9 to 807.1).

Next in Round 3 (conducted between September 20-24, 2020), we find a marginal improvement in weekly per capita expenditure across all the quintile classes as compared to Round 2, except for the top 20%. However, when compared to the week of February 2020, weekly per capita expenditure remains depressed across all the quintiles except for the top 20%. For example, for the bottom 20% consumption remain suppressed by -32.4% (95% CI: -39.2% to -25.3%). Similarly, for the top 60% to 80%, during the same period consumption remained depressed by -24.5% (95% CI: -33% to 16%). In contrast, for the top 20%, there is recovery in the weekly per capita expenditure from Round 2 to Round 3, and when compared to February, consumption remains marginally and insignificantly depressed by -8.2% (95% CI: -17.8% to 1.4%) (see figure 1).

Our next set of results focuses on consumption inequality by examining the consumption share of the percentile groups. In the week of February 2020, the share of the top 20% share in total consumption was 46.6% (95% CI: 44.8% to 48.5%), while that of the bottom 20% was 5.5% (95% CI: 5.1% to 5.9%). However, by Round 3 (in September 2020), the share of the top 20% went up to 52.3% (95% CI: 50.3% to 54.3%) - a significant increase of 5.7 percentage points (95% CI: 3% to 8.4%). On the other hand, the consumption share declined significantly for all the other quintile groups. For example, for the 60% to 80% quintile group, the share in total consumption fell from 22.5% (95% CI: 21.2% to 23.8%) to 20.7% (95% CI: 19.8% to 21.6%) - a decline of -1.7 percentage points (95% CI: -3.3% to -0.2%). (See table 1 and figure 1).

Finally, we look at the Gini coefficient of consumption distribution, and find that it increased from 0.407 in February 2020 (Round 1) to 0.469 in Round 3 (September 2020). (See Table 1 and Figure 1 for detailed results).

To sum up, the analysis of consumption expenditures across the three Rounds (February, July and September 2020) reveals that:

∘ in the early periods of the pandemic, and during the lockdown, consumption decline was most severe for the bottom 40% followed by the quintile class 40% to 60%. For the 60% to 80% quintile class, consumption declined insignificantly. For the top 20%, consumption increased marginally though insignificantly;
∘ as the pandemic deepened, there was a severe contraction in consumption across all quintile classes; and
∘ inequality in consumption expenditures (as captured by the Gini coefficient) increased over the three Rounds between July and September 2020

## 4. Discussion

COVID 19 pandemic is the worst economic disaster since the Great Depression.^8^ It is estimated that the pandemic cost the world economy a little more than USD 10 trillion in terms of foregone GDP in 2020-21.^9^ The IMF reckons that close to 90 million people might have been pushed into poverty.^10^ Unfortunately, the pandemic has not been a great leveler of income or consumption inequality.

India faces a fresh challenge of mitigating the crisis unleashed by the ‘second wave’ where the daily cases in May 2021 have been three times larger than in the first wave between March and September 2020. This has once again prompted several state governments to impose micro lockdowns to contain the virus. Unfortunately it has also unleashed an atmosphere of fear, panic, and uncertainty.^3,4^

The findings presented in this paper have significant implications for mitigating the adverse consequences of COVID-19. The first wave of the COVID 19 pandemic significantly reduced consumption across all the classes of people over time as compared to consumption prior to the pandemic. The reduction in consumption expenditures was not uniform either over time or across the classes. Two features, however, stand out:

∘ In the early phase of the pandemic, it was the bottom 40% that experienced a significant decline. As the pandemic deepened, consumption declined across all the classes, and was particularly severe for the middle class, the middle 40% to 80%.
∘ Consumption for the middle and the bottom class remained suppressed till September 2020, almost six months into the pandemic.

Early evidence indicates that people from disadvantaged socio-economic groups have had a much bigger economic fallout, particularly in terms of job losses, and severe health consequences compared to the more advantaged socio-economic groups.^6,10–13^ However, in the Indian context, it is not only the poor households but also the middle class, with limited support from the government, that has had to face the brunt of the crisis.

As we move forward, the Union and state governments would need to consider a new round of fiscal expansion to stimulate aggregate demand. It might not be enough for COVID-relief packages to target only the poor with, for instance, (a) the provision of an additional five kilograms of food grains and one kilogram of pulses to poor households under the National Food Security Act, (b) provision of free of cost LPG refills for gas cylinders for three months starting April 2020 to below-the-poverty-line households under the Pradhan Mantri Ujjwala Yojana, and (c) a cash transfer of INR 500 for the period April to June 2020 to women with Jan Dhan Yojana account holders.^6^

In addition to focusing on poor households, policy responses to alleviate people’s sufferings might have to consider a more comprehensive boost to consumption and compensate for the reduced consumption among middle class families as well.

Another important aspect to explore would be the impacts of the reductions in consumption levels on the nutritional security of households. According to the High Level Panel of Experts on Food Security and Nutrition, “…the virus, and measures to contain its spread, have had profound implications for food security, nutrition and food systems.” The COVID-19 pandemic is expected to increase the risk of all forms of malnutrition.^14^ Loss of household incomes, changes in the availability and affordability of nutritious foods, as well as disruptions caused to health and nutrition services by the lockdowns are expected to lead to an increase in child malnutrition, including wasting.^15^ The Government should consider more pro-active responses to offset the specific nutritional deficiencies that could arise out of the loss of incomes and declines in consumption following the COVID-19 pandemic.

Another short coming highlighted by the COVID-19 pandemic has been the poverty of data to make informed decisions. Rich countries that have invested in robust and periodic collection of conventional economic data, such as employment and consumption, are at an advantage to tailor their policies to the needs of their people, whereas low and middle income countries that either had no incentive to build a robust statistical system or had any use for them are at a big disadvantage to allocate resources to those who need them the most.^16^

India is one of the few low-middle income countries that has a robust statistical system and collects high frequency data on a variety of indicators such as consumer goods (durable and non-durable), manufacturing, and fuel consumption.^17^ However, the COVID-19 pandemic reveals that the available data are not very helpful in taking real time decisions for framing targeted policies to mitigate the losses suffered by different classes of people. There is urgent need to invest in statistical capabilities that collect real time data at least on consumption, unemployment, and health.

## 5. Limitations

A key limitation of the data used in this analysis is that the survey was conducted over phone with a relatively low response rate. This raises questions regarding the representativeness and external validity of the data. It is likely that poor households with limited or no access to phone could be underrepresented, implying higher estimates of declines in consumption for poor households than what the reality is.

## 6. Conclusion

Our paper highlights that the COVID-19 pandemic has, in the short run, suppressed consumption not only of the poor households but it significantly reduced consumption of the middle classes. Second, consumption inequalities have worsened as the consumption share of the top 20% increased, while it reduced significantly for all the other classes. Finally, the Central and state governments ought to take a more holistic account of the losses suffered by households to include the middle classes along with the poor as they prepare for another fiscal push to boost aggregate demand and counter the adverse effects of the second wave of the COVID-19 pandemic.

## Data Availability

India - COVID-19-Related Shocks in Rural India 2020, Rounds 1-3. https://microdata.worldbank.org/index.php/catalog/3830.

